# Intra- and interindividual variability in fasted gastric content volume

**DOI:** 10.1101/2024.03.12.24304085

**Authors:** Julia J.M. Roelofs, Guido Camps, Louise M. Leenders, Luca Marciani, Robin C. Spiller, Elise J.M. van Eijnatten, Jaber Alyami, Ruoxuan Deng, Daniela Freitas, Michael Grimm, Leila J. Karhunen, Shanthi Krishnasamy, Steven Le Feunteun, Dileep N. Lobo, Alan R. Mackie, Morwarid Mayar, Werner Weitschies, Paul A.M. Smeets

## Abstract

2

**Background:** Gastric fluid plays a key role in food digestion and drug dissolution, therefore, the amount of gastric fluid present in a fasted state may influence subsequent digestion and drug delivery. We aimed to describe intra- and interindividual variation in fasted gastric content volume (FGCV) and to determine the association with age, sex, and body size characteristics.

**Methods:** Data from 24 MRI studies measuring FGCV in healthy, mostly young individuals after an overnight fast were pooled. Analysis included 366 participants with a total of 870 measurements. Linear mixed model analysis was performed to calculate intra- and interindividual variability and to assess the effects of age, sex, weight, height, weight*height as a proxy for body size, and body mass index (BMI).

**Results:** FGCV ranged from 0 to 156 mL, with a mean (± SD) value of 33 ± 25 mL. The overall coefficient of variation within the study population was 75.6%, interindividual SD was 15 mL, and the intraindividual SD was 19 mL. Age, weight, height, weight*height, and BMI had no effect on FGCV. Women had lower volumes compared to men (MD: -6 mL), when corrected for the aforementioned factors.

**Conclusion:** FGCV is highly variable, with higher intraindividual compared to interindividual variability, indicating that FGCV is subject to day-to-day and within-day variation and is not a stable personal characteristic. This highlights the importance of considering FGCV when studying digestion and drug dissolution. Exact implications remain to be studied.

**GRAPHICAL ABSTRACT:** 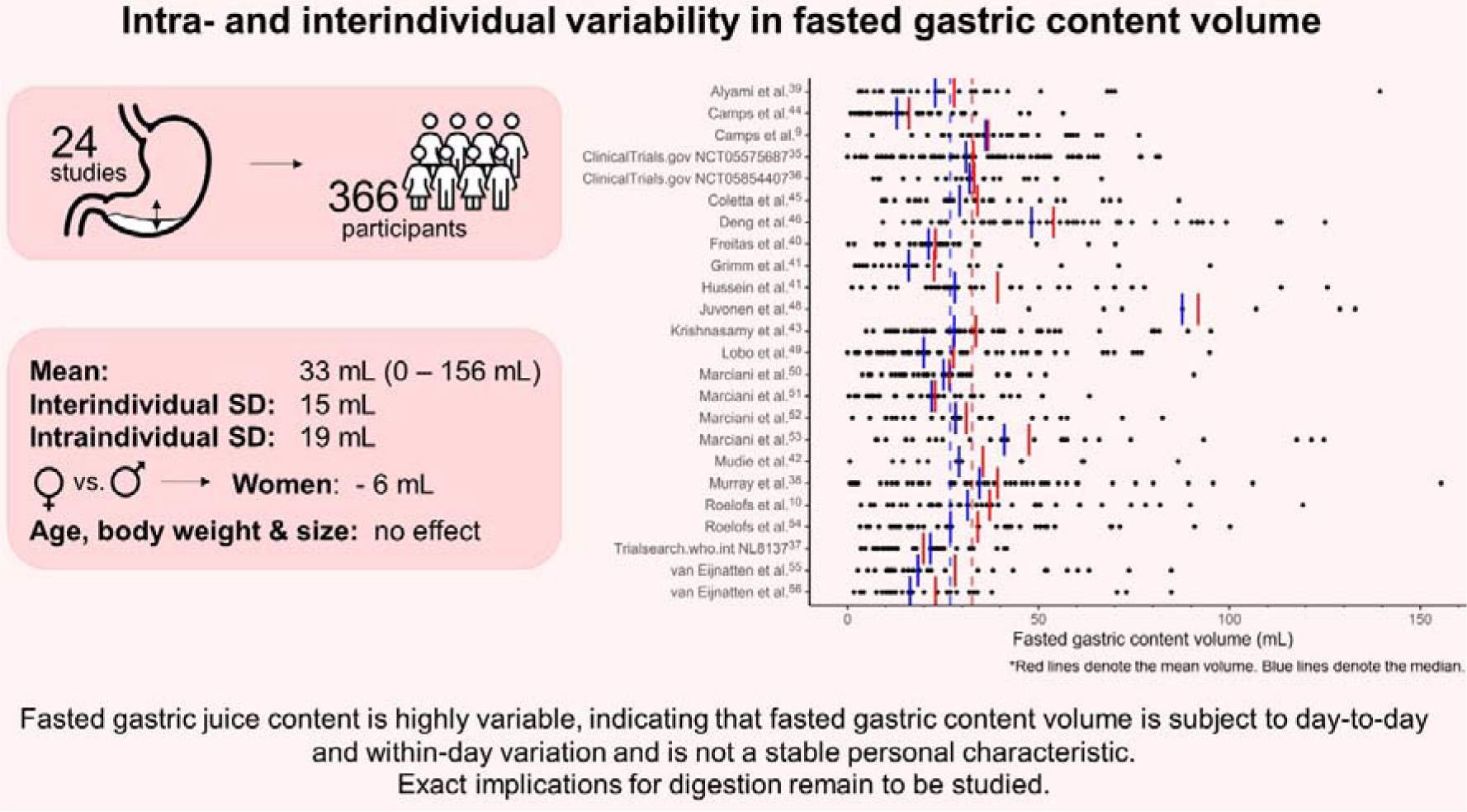

**KEY POINTS:**

- Fasted gastric content volume is highly variable, both within an individual and between individuals, and should range between 0 and 138 mL in healthy young individuals.
- Women have lower fasted gastric content volume compared to men; age, body weight and body size were not associated with differences in fasted gastric content volume.
- Fasted gastric content volume can impact both digestion and drug dissolution, although exact implications of the observed variations remain to be studied.

## 3 Introduction

Digestion is the breakdown of food into particles that can be absorbed by the body. This process starts with the oral phase, where mastication and secretion of saliva lead to the formation of a food bolus that can be swallowed safely, and continues in the gastrointestinal tract^1,2^. Digestion is a series of mechanical, physiological, and biochemical processing steps that eventually allows for absorption and utilization of nutrients^3^. These biochemical processing steps include the breakdown by acid and enzymes present in gastric secretions. Gastric fluid serves two main functions: it acts as a first line of defense against infection by killing swallowed microorganisms, and it aids in digestion by initiating the breakdown of food^4^. Gastric fluid is a combination of water, hydrochloric acid (HCl), gastric lipase, pepsin, intrinsic factor, ions (Na^+^, K^+^, and Cl^-^), and mucus^4,5^. These components are continuously secreted by endocrine cells in the stomach wall to maintain an acidic environment with a pH between 1.4 and 2.0 in the fasted state^5,6^. The fasted gastric secretion rate is ∼1 mL min^-1^ but after food ingestion it can increase to up to 9 mL min^-1^ ^6,7^.

The main zymogen in gastric fluid is pepsinogen^7,8^. Under acidic conditions, pepsinogen converts into its active form, pepsin^5,7^. A pH ≤ 2.0 allows pepsin to function optimally, while a pH > 7.2 irreversibly denatures it. The secretion of HCl is therefore essential for the activity of pepsin^8^. Pepsin is important for the digestion of proteins as it breaks them down into smaller peptides^7^. Due to its role in protein digestion, the amount of gastric fluid present in the fasted stomach will influence gastric protein digestion by affecting the pH and amount of pepsin available. Camps et al.^9^ found that FGCV affected gastric layering and gastric emptying of infant formula. In line with this, Roelofs et al.^10^ found that FGCV was highly correlated with the destabilization of infant formula in the stomach: a higher FGCV correlated with earlier gastric phase separation of the emulsion. Since the emulsion was stabilized with casein-whey complexes, this effect is likely explained by increased protein hydrolysis and flocculation due to the low pH and the presence of pepsin in the stomach. When food is ingested, gastric pH increases due to the buffering effect of the meal^11^. However, with a higher FGCV, the gastric pH will initially be lower for a given buffering effect. In addition, more pepsin will also be available. Since pepsin activity is higher at low pH^12^, a higher fasted gastric fluid volume is thus associated to both a higher amount of pepsin and a higher pepsin activity, leading to increased protein hydrolysis in the early stages of digestion^6^. Moreover, FGCV is also a crucial parameter in oral drug delivery. Changes in fasting volumes and composition contribute to intra- and interindividual variability in drug plasma profiles^13,14^. Data on the variability are therefore much needed in order to optimize *in vitro* and *in silico* models for the development of novel drugs and dosage forms^15^.

Many studies that investigated variability in fasted gastric fluid have used gastric aspiration to measure the stomach content. Gastric aspirates are taken through a nasogastric- or endoscopic tube and is usually done over a 15 to 60-minute period. Nasogastric intubation is an invasive procedure which could perturb the baseline physiological state and the progression of the tube from the throat to the stomach is often aided with water swallows. Gastric emptying also depends on the actual positioning of the tube ports within the stomach. Although total gastric content can be measured with this method, results are often reported as secretion rates^16^. More recently, magnetic resonance imaging (MRI) has been used to measure gastric content volume. This method is less invasive and the technique is inherently suited to visualize body fluids with excellent spatial resolution. In contrast to gastric aspirates, the use of MRI yields measurement of gastric volume at specific time points.

Other studies found that various individual characteristics may affect gastric acid secretion. Goldschmiedt et al.^17^ found a trend towards an association of aging with gastric acid secretion and a higher basal acid output in older (44-71 y) compared to younger (23-42 y) adults (5.8 vs. 3.2 mmol h^-1^, p = 0.05). When the sexes were tested separately, older men (45-71 y) had significantly higher acid outputs than younger men (27-37 y, output: 8.4 vs. 3.8 mmol h^-1^). Older women (44-65 y) tended to have slightly higher acid outputs than younger women (23-42 y, p = 0.09) and older men had a higher output compared to older women. In contrast, Feldman et al.^18^ found no difference in fasted gastric acid secretion between young (18-34 y), middle-aged (35-64 y) and elderly (65-98 y). Studies showed that, on average, women have lower fasted gastric acid secretion rates compared to men. Feldman et al.^19^ found that basal acid output was almost twice as high in men compared to women. It has been suggested that this is due to hormones, lower body weight or smaller stature of women, thus having smaller stomachs, and associated decreased parietal cell mass^19-25^. Moreover, body weight was found to be weakly correlated with fasted gastric acid secretion rate in young adults (n = 176, r = 0.184)^26^. Altogether, these findings suggest a possible role of age, sex, and body size characteristics on FGCV. However, these studies all reported gastric acid secretion rate as measured by gastric aspiration as opposed to gastric fluid secretion rate or gastric volume. Moreover, taking gastric aspirates might result in underestimation as some of the gastric acid might be lost due to gastric emptying. In addition, removing the gastric juice from the stomach itself can reinforce the secretion^27^. Furthermore, the migrating motor complex (MCC) cycle is known to cause temporal changes in gastric motility and secretion^13^ and might therefore contribute to the variation in FGCV.

Grimm et al.^15^ compared FGCV from 5 MRI studies with 1-6 visits. They found a mean fasted volume of 25 ± 18 mL (n = 120), with a range from 1 to 96 mL. The interindividual and intraindividual variability were comparable, namely 49 ± 19 mL and 44 ± 18 mL, respectively.

To date, several MRI studies have reported FGCV. However, the sample sizes in these studies are generally small. The aim of this study was therefore to gather all available data and combine it to provide unique insights on the intra- and interindividual variation in FGCV and to explore possible associations with age, sex, and body size characteristics.

## 4 Methods

### 4.1 Study selection

Studies were selected by contacting members of the INFOGEST and UNGAP Imaging Working Group^28^ for available data and by emailing authors of eligible studies. A PubMed search was performed in May 2023 to identify eligible studies using the following search string: (all fields): “(gastric AND (emptying OR retention) AND ("Magnetic Resonance Imaging" OR MRI) [full text, clinical trial, human, English]”. Inclusion criteria for studies were: 1) data on FGCV was determined from MRI images, 2) the study was published in a peer reviewed journal and/or registered in a public trial registry, 3) the study was published in the past 15 years (in or after 2008) and 4) the study was conducted in healthy participants. Studies that did not include a fast of at least 10 h prior to the assessment of FGCV were excluded.

All volunteers provided written informed consent for the specific study procedures and subsequent use of anonymized volume data for comparative investigations. Authors who agreed to participate provided data on FGCV and individual participant characteristics (sex, age, weight, and height). Information on in- and exclusion criteria was extracted from the paper or the trial registration. This analysis was preregistered at OSF under code BDQS4 (https://doi.org/10.17605/OSF.IO/BDQS4).

### 4.2 Statistical analysis

FGCV data was characterized by means, medians, standard deviation, range, and overall coefficient of variation. A linear mixed model was used to assess intra- and interindividual variability and the effects of age, sex, and body size characteristics. Participant ID and study site were added as random factors. Age, sex, weight, height, weight*height, and BMI were added as fixed factors in two different models. That is, a model with age, sex, weight and height, and a model with age, sex, weight*height, and BMI. Non-collinearity of the variables was confirmed in both models with the Variance Inflation Factor (VIF) (all < 3)^29^. In addition, the intraindividual range (maximum value – minimum value) was calculated for each person to be able to compare our results to the study of Grimm et al.^15^.

To determine what a normal range of variation in FGCV might be, we calculated mean ± 3 SDs, which should cover 99.7% of the data. Since our data on gastric content volumes followed a right-skewed distribution, we applied this method to our square root transformed data and back transformed the outcome.

Normality of the data was assessed with quantile-quantile (QQ) plots of the residuals, which showed a roughly normal distribution. Using a square root transformation led to a slight improvement in the distribution but had no effect on our outcomes. Transforming the data leads to difficulty in interpreting the data, and because linear mixed models are capable of dealing with violation of the distributional assumptions^30^, we chose to report the results of the non-transformed data. Analysis was performed in R version 4.1.3, using the nlme^31^ and car package^32^. The significance threshold was set at p = 0.05. Data are expressed as mean ± SD unless stated otherwise.

## 5 Results

### 5.1 Included studies

In total, data from 31 studies was received. Four studies were excluded due to a fasting period of less than 10h and two due to missing data. Data of four unpublished studies were received^33-35^ via the INFOGEST and UNGAP Imaging Working Group^28^, of which one was excluded due to unavailability of a trial registration. This led to inclusion of 24 studies in total, performed at 5 research sites in the UK, Germany, France, and the Netherlands (**Figure 1**).

**Figure 1.**
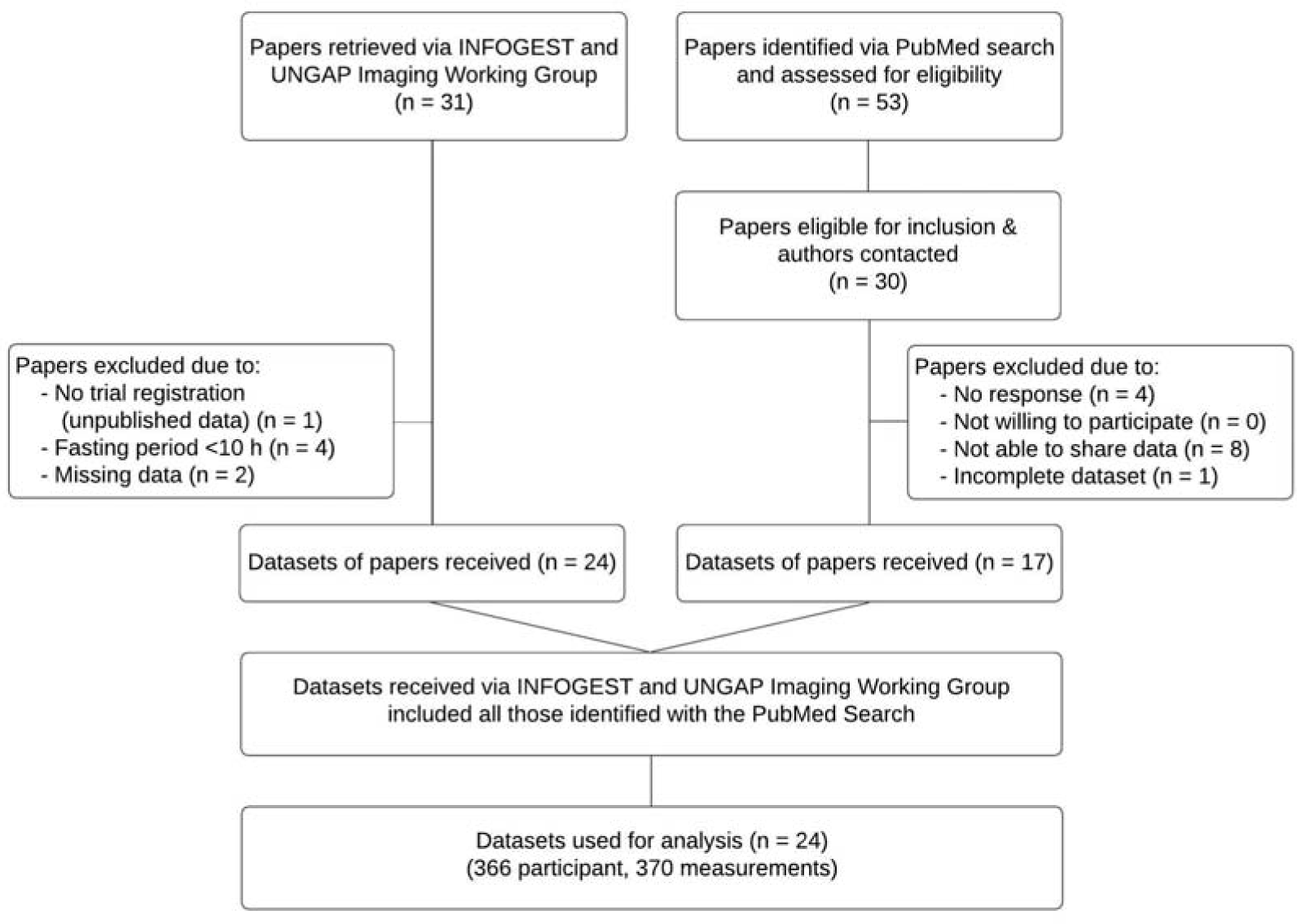
Flow chart.

### 5.2 Study designs

The fasting regimen differed somewhat between the studies; details can be found in **Supplementary Table 1**. In short, all studies included an overnight fast of at least 10 hours (the selection criterion). In some studies, participants were not allowed to drink during the fast, while others could drink water (or non-caloric, non-caffeinated drinks) up until 1-2 hours before their visit. Five studies standardized the evening meal before the fasting. Fourteen studies included instructions on physical activity, either not allowing heavy exercise or keeping it constant before and on all test days. Studies that did not exclude participants based on medication use required the participants to quit the medication for at least the duration of the overnight fast.

Inclusion and exclusion criteria differed slightly between studies. Details can be found in Supplementary Table 2.

### 5.3 MRI measurements

All studies scanned the participants during a breath hold to fixate the position of the diaphragm and the stomach and prevent respiratory motion artefacts. Most studies scanned participants in a supine position, but two scanned participants in a supine tilted position with their left side slightly raised^36,37^ and one scanned participants in a right decubitus position^38^. This was not expected to influence gastric secretion, however, no literature could be identified on this. A study comparing gastric emptying of a soup in the right versus left decubitus position found no difference^39^.

Since the studies were performed at different research facilities, scanning protocols slightly deviated from each other. However, this is not expected to influence the results. Details of the scan sequences used can be found in **Supplemental Table 3**. All studies acquired transverse slices, except for Grimm et al.^40^ who acquired coronal slices.

Three studies analyzed gastric content volumes semiautomatically using an intensity-based thresholding technique^40-42^. All other studies manually delineated the stomach contents on each slice of the MRI scan to calculate the total gastric content volume.

### 5.4 Participants

In total, 366 participants were included in the analysis, 146 women and 220 men. Participants were 25.5 ± 6.8 years old and had a BMI of 22.7 ± 2.3 kg m^-2^ (**Table 1 & 2**). In total, 870 individual measurements were collected, of which one measurement was considered an outlier and excluded for analysis. This participant had a baseline FGCV volume of 373 mL, which is 12.3 SDs above the mean, which seems highly unlikely caused by natural variation. In addition, the same participant showed a FGCV of 21 and 26 mL on other days. Therefore, this high volume of 373 mL was deemed unrealistic. It might have been caused by non-compliance to the fasting period or a rare case of gastroparesis. The analysis was performed both on the complete dataset, as well as the set without outlier. Results of the analysis with the outlier are reported in **Supplementary Tables 4 & 5**.

**Table 1.** Overview of studies and their participant characteristics.

| Study | Sex<br>(# woman/men) | Age<br>(years) | BMI<br>(kg m<sup>-2</sup>) | # measurements<br>per participant |
| --- | --- | --- | --- | --- |
| Alyami et al.<sup>37</sup> | 13/6 | 28.2 ± 10.6 | 23.4 ± 3.1 | 2 |
| Camps et al.<sup>43</sup> | 0/19 | - | 21.7 ± 1.4 | 2 |
| Camps et al.<sup>9</sup> | 16/0 | 30.9 ± 3.7 | 22.6 ± 3.9 | 2 |
| ClinicalTrials.gov NCT05575687<sup>33</sup> | 6/6 | 24.1 ± 4.3 | 22.2 ± 2.2 | 2 |
| ClinicalTrials.gov NCT05854407<sup>34</sup> | 10/6 | 22.8 ± 1.8 | 22.1 ± 2.0 | 6 |
| Coletta et al.<sup>44</sup> | 7/5 | 27.3 ± 8.4 | 22.7 ± 1.8 | 3 |
| Deng et al.<sup>45</sup> | 0/18 | 27.1 ± 4.9 | 22.8 ± 1.6 | 3 |
| Freitas et al.<sup>38</sup> | 0/10 | 32.8 ± 10.4 | 23.0 ± 1.8 | 3 |
| Grimm et al.<sup>40</sup> | 3/3 | 25.2 ± 2.1 | 23.4 ± 1.2 | 4 |
| Hussein et al.<sup>46</sup> | 0/11 | 23.8 ± 4.1 | 24.4 ± 3.2 | 3 |
| Juvonen et al.<sup>47</sup> | 0/4 | 39.0 ± 8.0 | 25.2 ± 1.0 | 2 |
| Krishnasamy et al.<sup>42</sup> | 14/4 | 24.7 ± 4.2 | 22.7 ± 3.5 | 3 |
| Lobo et al.<sup>48</sup> | 10/10 | 29.4 ± 7.8 | 23.4 ± 1.9 | 2 |
| Marciani et al.<sup>49</sup> | 8/8 | 22.8 ± 3.7 | 22.0 ± 2.3 | 2 |
| Marciani et al.<sup>50</sup> | 9/9 | 20.3 ± 0.8 | 22.2 ± 2.2 | 2 |
| Marciani et al.<sup>51</sup> | 4/8 | 22.3 ± 4.4 | 23.0 ± 2.9 | 2 |
| Marciani et al.<sup>52</sup> | 5/8 | 20.8 ± 0.7 | 22.6 ± 1.7 | 2 |
| Mudie et al.<sup>41</sup> | 8/4 | 21.3 ± 2.2 | 22.1 ± 2.0 | 1 |
| Murray et al.<sup>36</sup> | 0/17 | 25.3 ± 5.5 | 23.8 ± 2.7 | 3 |
| Roelofs et al.<sup>10</sup> | 0/20 | 25.5 ± 5.8 | 21.9 ± 1.5 | 2 |
| Roelofs et al.<sup>53</sup> | 0/14 | 23.0 ± 3.8 | 22.2 ± 1.7 | 3 |
| Trialsearch.who.int NL8137<sup>35</sup> | 0/18 | 25.9 ± 8.3 | 22.7 ± 1.6 | 2 |
| van Eijnatten et al.<sup>54</sup> | 0/15 | 30.9 ± 13.8 | 22.3 ± 2.0 | 2 |
| van Eijnatten et al.<sup>55</sup> | 30/0 | 25.1 ± 5.2 | 22.4 ± 2.3 | 1 |
| **Total** | **146/220** | **25.5 ± 6.8** | **22.7 ± 2.3** |  |

**Table 2.** Mean ± SD age and body mass index (BMI) of study participants, stratified by sex.

|  | **Women**<br>**(n = 146)** |  |  | **Men**<br>**(n = 220)** |  |  | **Total**<br>**(n = 366)** |  |  |
| --- | --- | --- | --- | --- | --- | --- | --- | --- | --- |
|  | *Mean ± SD* | *Median* | *Range* | *Mean ± SD* | *Median* | *Range* | *Mean ± SD* | *Median* | *Range* |
| **Age (y)** | 24.9 ± 6.0 | 23 | 18-57 | 25.9 ± 7.2 | 23 | 18-55 | 25.5 ± 6.8 | 23 | 18-57 |
| **BMI (kg m<sup>-2</sup>)** | 22.2 ± 2.2 | 22.1 | 18.0-30.4 | 23.0 ± 2.3 | 22.7 | 18.3-33.0 | 22.7 ± 2.3 | 22.5 | 18.0-33.0 |
| **Weight (kg)** | 61.9 ± 7.8 | 62.5 | 44.0-90.0 | 75.4 ± 9.4 | 74.0 | 58.0-108.0 | 70.4 ± 11.0 | 70.0 | 44.0-108.0 |
| **Height (m)** | 1.67 ± 0.07 | 1.67 | 1.49-1.90 | 1.81 ± 0.08 | 1.80 | 1.54-2.02 | 1.76 ± 0.1 | 1.77 | 1.49-2.02 |
| **Height\*Weight (m\*kg)** | 103.6 ± 16.2 | 103.2 | 66.0-156.6 | 136.9 ± 21.2 | 134.0 | 89.3-199.0 | 124.6 ± 25.3 | 123.7 | 66.0-199.0 |
| **Fasted gastric content volume (mL)** | 31 ± 21 | 27 | 0.0-122 | 33 ± 27 | 27 | 0-156 | 33 ± 25 | 27 | 0-156 |

The studies included up to six measurements of FGCV per participant, with the majority of the participants having two or three measurements. There were 47 participants with one measurement, 184 with two, 115 with three, 6 with four, and 14 with six measurements.

### 5.5 Fasted gastric content volume

FGCV ranged from 0 mL to 156 mL, with a mean value of 33 ± 25 mL (**Figure 2**, **Table 2**). An example of a relatively low, medium and high FGCV is shown in **Figure 3**. The median volume was 27 mL (IQR = 15 – 45 mL). Women had a slightly lower FGCV compared to men (31 ± 21 mL vs. 33 ± 27 mL). The overall coefficient of variation within the study population was 75.6%. The interindividual standard deviation, corrected for study site, was 15 mL, while the intraindividual standard deviation was 19 mL. The intraindividual range varied between 0 and 108 mL, with a mean of 24 mL and a SD of 21 mL.

**Figure 2.**
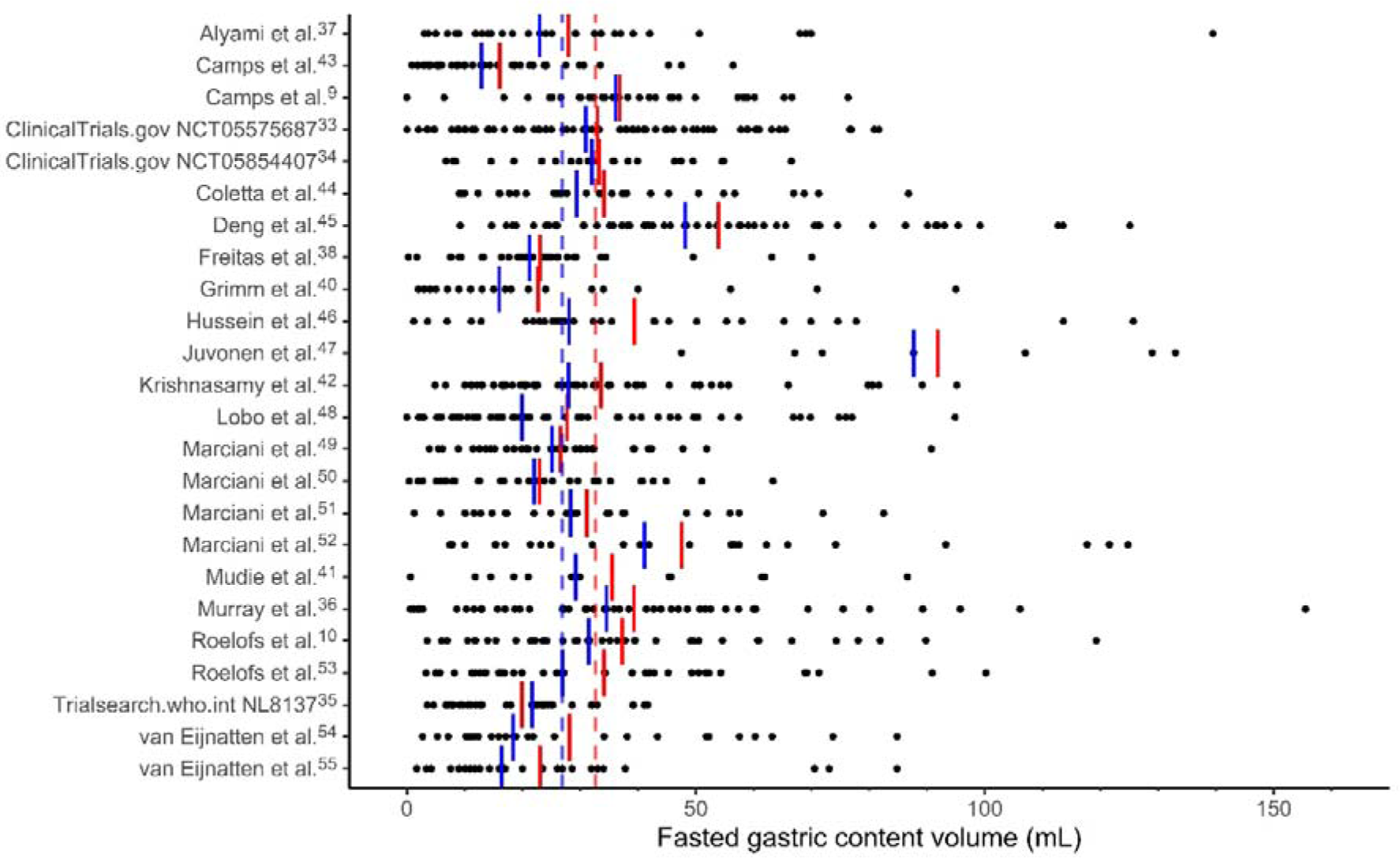
Fasted gastric content volumes for each study and for all individual visits (black dots). Red lines denote the mean volume. Blue lines denote the median. The dashed lines denote the overall mean (red) and median (blue).

**Figure 3.**
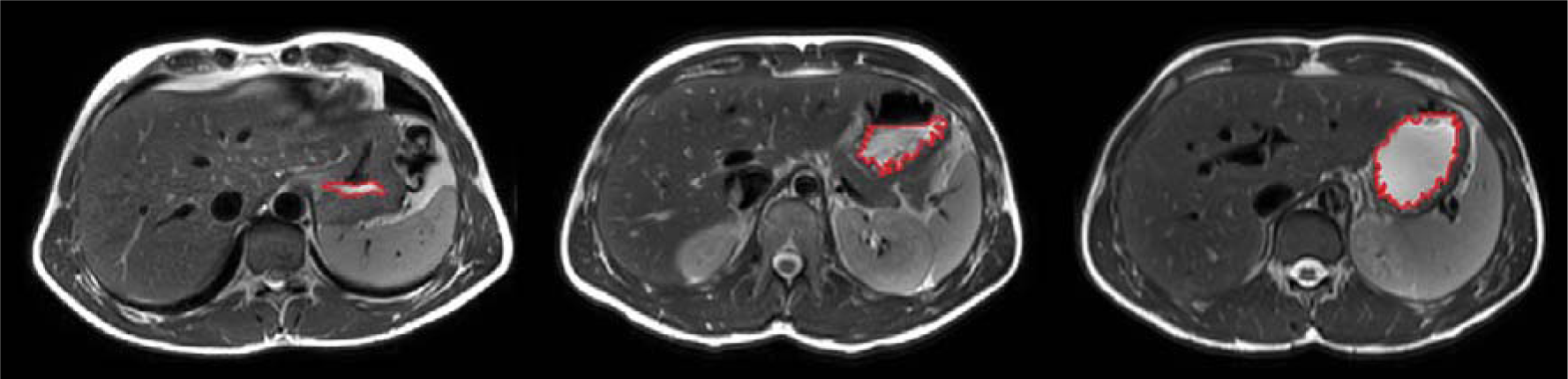
Illustration of fasted gastric content volume showing MRI images for three different volumes (left to right: 4 mL, 61 mL, and 119 mL).

Calculating the mean ± 3 SD, results in a FGCV that would range between 0-138 mL. This is in fair agreement with the volumes in our data, only 2 measurements are outside of this range (0.2% of the values).

### 5.6 Age, sex, and body size characteristics

There was no effect of age, weight, height, weight*height, and BMI on FGCV (**Table 3**). For both models, men showed a significantly higher volume compared to women (estimate: -6 mL, p = 0.045 and p = 0.043, for the model with weight & height and the model with weight*height and BMI, respectively). Scatterplots of age, sex, and BMI with FGCV are shown in **Figure 4**, those of weight, height, and weight*height can be found in **Supplementary Figure 2**.

**Figure 4.**
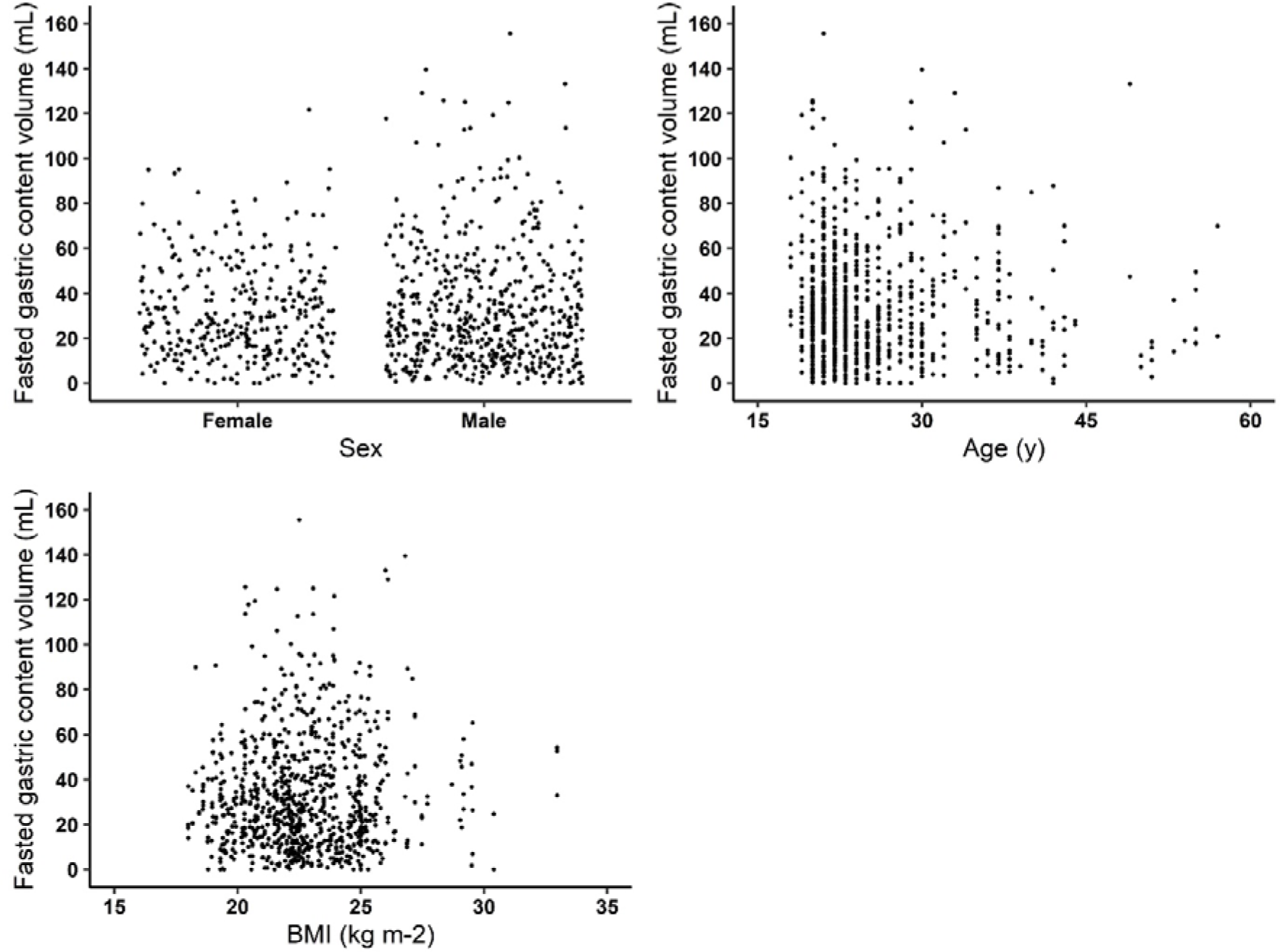
Scatterplots of fasted gastric content volume by age, sex, and body mass index (BMI). Linear mixed model analysis showed that women had lower fasted gastric content volume compared to men (∼6 mL, p < 0.05). There was no effect for age and BMI.

**Table 3.**
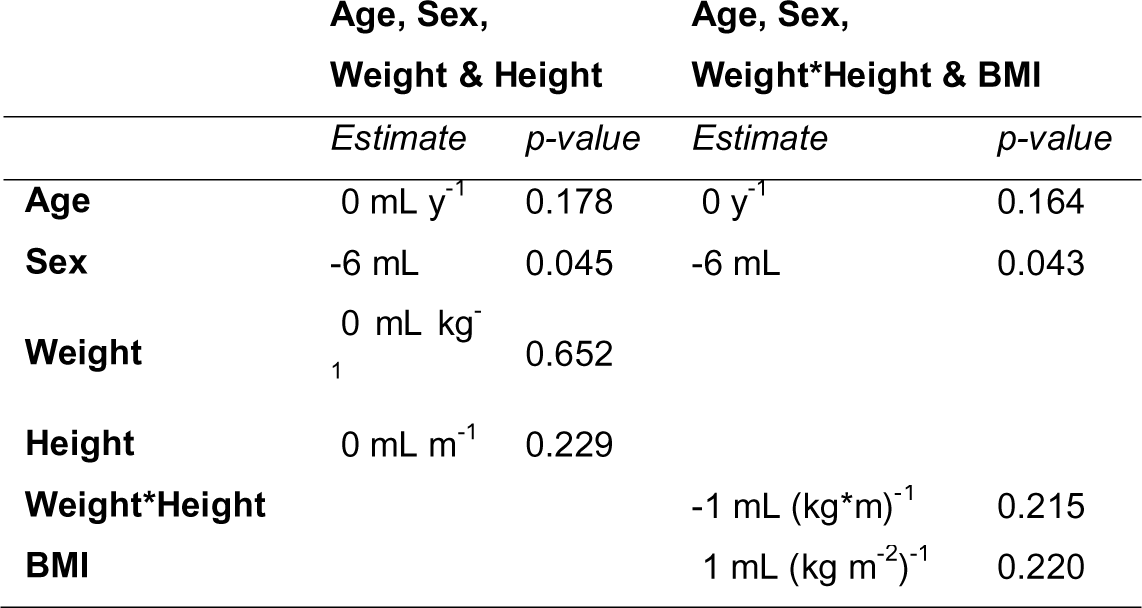
Results of the linear mixed model analysis of the effect of age, sex, and body size characteristics on fasted gastric content volume (mL unit^-1^).

|  | Age, Sex,<br>Weight & Height |  | Age, Sex,<br>Weight*Height & BMI |  |
| --- | --- | --- | --- | --- |
|  | *Estimate* | *p-value* | *Estimate* | *p-value* |
| **Age** | 0 mL y<sup>-1</sup> | 0.178 | 0 y<sup>-1</sup> | 0.164 |
| **Sex** | -6 mL | 0.045 | -6 mL | 0.043 |
| **Weight** | 0 mL kg<sup>-1</sup> | 0.652 |  |  |
| **Height** | 0 mL m<sup>-1</sup> | 0.229 |  |  |
| **Weight\*Height** |  |  | -1 mL (kg*m)<sup>-1</sup> | 0.215 |
| **BMI** |  |  | 1 mL (kg m<sup>-2</sup>)<sup>-1</sup> | 0.220 |

## 6 Discussion

With this analysis we aimed to establish the intra- and interindividual variation in FGCV, and to assess to what extent this variation is associated with age, sex, and body size. Our analysis, based on 366 participants, established that there is large variation in FGCV within healthy adults, with a mean of 33 mL and overall coefficient of variation of 75.6%. When corrected for age, sex, and body size, interindividual variability was 15 mL and intraindividual variability 19 mL. Age, weight, height, weight*height, and BMI were not associated with FGCV. Men were found to have a ∼6 mL higher FGCV compared to women, although clinical relevance remains to be studied.

### 6.1 Intra- and interindividual variability

The mean FGCV of 33 ± 25 mL is comparable to the mean volume of 25 ± 18 mL (n = 120) found by Grimm et al.^15^. Their volumes ranged from 1 to 96 mL, while ours ranged up until 156 mL. Although our upper limit is more than 1.5 times as high, only 1.8% of our volumes were above 100 mL (i.e., 16 out of 869) indicating that the majority of our data was within the same range as theirs. Based on our results, a normal FGCV can range from 0-138 mL. The coefficient of variation was 75.6%. Data in literature is limited, Grimm et al.^15^ reported values ranging from 39 to 159%, although sample size was very small (n = 6 each). Our mean intraindividual range was 24 ± 21 mL, which is much lower compared to that of Grimm et al.^15^ of 44 ± 18 mL but might be explained by their low sample size (n = 8 subjects), which is more susceptible to individuals with high variability.

Interindividual variation was slightly lower with 15 mL compared to the intraindividual variation of 19 mL. This indicates that FGCV is subject to day-to-day and within-day variation and cannot be seen as a stable personal characteristic. Part of the variation might be explained by the MMC cycle. Studies have shown that the MMC causes temporal variations in gastric secretion. The different phases of the MMC cycle are associated with increases and decreases in secretion rate^13^ with differences up to 78%^56^. Therefore, the variability in both intra- and interindividual FGCV might partly be explained by the phase of the MMC cycle at the time of measurement. However, a review of van den Abeele et al.^13^ found that MMC cycle durations varied between 96 and 172 minutes and Parkman et al.^57^ found that only 1 in 3 individuals had antral contractions during a 60-minute period. Moreover, Kellow et al.^58^ reported that while MMC cycles occurred every 1-2 hours, only 1 in 3 originated in the stomach. Goetze et al.^59^ used MRI to repeatedly measure FGCV over 90 minutes but found no changes over this period (n = 12, average slope: 0.0018 mL min^-1^, p > 0.05)^59^. In addition to gastric secretion rates, the peristaltic, phasic contractions also change during the MMC cycle, resulting in an increased liquid emptying rate during phase III^13^. This increase in gastric secretion rate happens prior to phase III contractions, which roughly corresponds to late phase II/phase III contractions in the stomach^13^. This might therefore (in part) counteract the effect of the increased secretions during this phase. As gastric volume is the result of gastric secretion and emptying, this might explain why Goetze et al.^59^ found no difference in gastric volume over time. Altogether this indicates that more prolonged studies are needed to better capture the change in FGCV over time during the MMC cycle.

Comparison of our results with literature is difficult. Previous studies often report fasted gastric acid secretion rate as measured by gastric aspiration as opposed to gastric fluid secretion rate or gastric volume. The gastric aspirates are often taken over a period of 30-60 minutes and might therefore capture temporal fluctuations. A disadvantage is that this method might result in underestimation as some of the gastric acid might be lost due to gastric emptying. Moreover, removing the gastric juice from the stomach itself can reinforce the secretion^27^. This difference in assessment might account for some of the discrepancies between our findings and those in literature. One study has been identified that reported both volume and secretion rate. Goyal et al.^60^ collected gastric fluids over a period of 60-minute during continuous suction of gastric fluid. They found a mean volume of 63.5 mL and a gastric acid secretion rate of 2.99 mmol h^-1^. It is important to note that they urged the participants to spit out their saliva during collection. This is in contrast to measurements with MRI where the stomach might also contain small amounts of saliva. Saliva flow rate during fasting (6 hours) is 0.1 mL min^-1^, so minimal effects are expected^61^.

### 6.2 Biological variations and other factors influencing fasted gastric content volume

There was no effect of age on FGCV (-0.2 mL, p > 0.05), although it should be noted that our study population was relatively young, ranging 18-57 y with only 10 participants (2.7%) aged 44 years or older. Literature is inconclusive on the effect of age. Feldman et al.^18^ found no difference between young (18-34 y), middle-aged (35-64 y) and elderly (65-98 y) while Goldschmiedt et al.^17^ found a trend for higher FGCV in older (44-71 y) compared to younger (23-42 y) adults (5.8 vs. 3.2 mmol h^-1^, p = 0.05). Based on our findings, it can be concluded that within healthy, mainly young adults, age has no effect on FGCV when corrected for age, sex, and body size characteristics.

FGCV was lower for women compared to men (∼6 mL, p < 0.05), which is in agreement with previous studies that found higher fasted gastric secretion rates in men compared to women. It has been suggested that this might be due to their lower body weight or smaller stature, thus having smaller stomachs, and associated decreased parietal cell mass or due to hormonal differences^19-25^. Since we corrected for weight and BMI in our models, this could not have been the explanation for the difference between men and women. Novis et al.^26^ found that body weight was weakly correlated with fasted gastric acid secretion rate (n = 176, range: 45-105 kg, r = 0.184), however, we did not find an effect of either weight or BMI. Since it is known that the volume present in the stomach is the result of both secretion and gastric emptying and that those with a higher body weight have faster gastric emptying^62^, these effects might counterbalance the increase in secretion. Thus, based on our findings it can be concluded that weight, within a healthy range, does not influence FGCV.

In an attempt to correct for differences in stomach size, height and weight*height were included as a proxy for body size. No effect was found for these measures. It is, however, noteworthy that literature is inconclusive on whether stomach size is associated with weight and height. A study looking at mucosal surface area of the stomach *post-mortem* found that, on average, men have a 10% larger stomach size compared to women. However, variations in stomach size were not related to age, body height or weight^63^. In contrast, Lee et al.^64^ found that sex, age, height and body weight were associated with the length of the lesser curvature, and sex and weight with the length of the greater curvature. They found that men have longer stomachs compared to women, although it is noteworthy that this study was performed on stomachs removed during total gastrectomy in gastric cancer patients. Moreover, these studies included many overweight participants (>50%), raising the question whether these findings would be similar for healthy participants. Thus, although we corrected for body size by using height and weight*height, this might not have been very accurate for controlling for differences in stomach size. Whether stomach size, and associated parietal cell mass, is (partly) responsible for lower FGCV in women remains to be studied.

In addition to body size, hormones have been suggested as an explanation for sex differences in gastric acid secretion. Studies on the effect of sex hormones on gastric acid secretion are inconclusive. Goldschmiedt et al.^17^ did not find an effect of menstrual phase on fasted gastric acid secretion rate (n = 10). However, Sakaguchi et al.^65^ showed that gastric acid secretion is decreased in the premenstrual phase, showing an inverse correlation with plasma estradiol concentrations (n = 24, r = 0.629). An animal study showed that estrogen might inhibit gastric acid secretion by binding to estrogen receptors on the parietal cells in the stomach^66^. These findings might also explain the lower gastric acid secretion in women compared to men^65,66^. This effect of menstrual phase is something that has not been taken into account by all of the studies included in this analysis and might thus have contributed to the variation in fasting gastric volumes.

Gastric acid secretion is influenced by many more factors. Examples include, sex hormones, the circadian rhythm, smoking, exercise, stress levels, the use of medication, and the presence of *Helicobacter pylori*. Gastric acid secretion has been shown to have a distinct circadian cycle in the absence of food stimulation, with the highest secretion rate in the evening and the lowest in the morning^67^. Smokers have higher fasted gastric acid secretion compared to non-smokers (4.1 vs. 2.7 mmol h^-1^, respectively)^18^. Interestingly, not all studies mentioned the exclusion of smokers. Since the prevalence of smoking is higher among men^68^, this might have contributed to the difference we found between women and men. However, no data on the prevalence of smoking was available for the studies.

Studies on exercise and gastric acid secretion report a decrease in fasted gastric acid secretion either during exercise or during restitution^69-71^. Furthermore, physical stress is known to induce a 3-fold increase in fasted gastric acid secretion^72^. Moreover, certain medications are known to influence gastric acid secretion, such as protein pump inhibitors, antacids, and histamine, which might impact further digestion. For example, protein pump inhibitors are known to slow down the gastric emptying of solids, which is suggested to be due to impairment of intragastric peptic digestion^73^. Moreover, a review of Maideen^74^ found that long-term use of protein pump inhibitors is associated with micronutrient deficiencies (e.g. iron, B12, calcium).

Most of these factors are not expected to have influenced our results. Studies were all performed in the morning after an overnight fast, thereby minimizing the influence of the circadian rhythm and food and beverages that were consumed previously. In addition, the use of medication was either an exclusion criteria or stopped for at least the fasting period.

Moreover, it is known that a *Helicobacter pylori* infection can initially cause hypergastrinemia and gastric hypersecretion, while later in life it can cause gastric atrophy with impaired gastric secretion^75^. Since an infection is commonly asymptomatic, prevalence in Europe is around 34%^76^, and participants were not tested for this, this might explain some of the variance that we found.

### 6.3 Fasted gastric content volume and digestion

Gastric secretion is of key importance for (peptic) digestion. Naturally, the volume present in the stomach will affect digestion due to the presence of enzymes and HCl and their effect on the breakdown of nutrients, specifically proteins. Greater amounts of enzymes present and a higher concentration of HCl can both facilitate digestion in the early stages of gastric digestion. The association between FGCV and protein digestion was already shown^9,10^, where higher volumes were associated with earlier destabilization of emulsions, indicating increased protein hydrolysis. However, the sole effect of FGCV on digestion is difficult to establish given the many variables that are involved. Whether the difference in FGCV can be considered as clinically relevant also depends on the size of a meal: the larger the meal, the smaller the effects will likely be. Moreover, it not only depends on the initial amount of gastric juice present, but also on the increase in secretion that happens when food is ingested. This increase depends on multiple factors and can already start by the sight and smell of food, and increases further when tasting and chewing the food^77^. After that, the presence of the food and distention of the stomach will further stimulate gastric acid secretion. It was found that gastric secretion tends to increase with meal size. Furthermore, the constituents of food can affect the gastric acid secretion, e.g. it is known that peptides and amino acids stimulate secretion, but also alcohol^78,79^, calcium^5^, and lemon juice^38^

Limited literature is available on the correlation between fasting and meal-induced gastric secretion. Goyal et al.^60^ found no correlation between basal and histamine-stimulated peak total secretion (r = 0.345, n = 22). Moreover, they reported ratios of 4.7-45.3% between basal and histamine-stimulated maximal acid output (n = 22), with all, except one, below 35%. Other studies found ratios between 9.9 and 30.7%^60^. Thus, based on FGCV alone, it is difficult to predict the exact effect on digestion. This highlights the need for methods to estimate gastric acid secretion after ingestion of food. Marciani et al.^80^ used T_2_ relaxation time measurements to monitor the process of dilution by gastric secretions and mixing of viscous meals and Goetze et al.^59^ used fast T_1_ mapping techniques for the quantification of intra-gastric dilution and distribution of orally applied gadolinium-based paramagnetic contrast agents showing that there is potential for estimating gastric secretion volumes with MRI.

Moreover, it is important to question what volume can be considered as a clinically relevant difference in FGCV and will impact digestion kinetics. Recently, an *in vitro* digestion model was developed that considers sex differences in the gastrointestinal tract, accounting for the lower fasting and meal-stimulated gastric acid secretion rates and higher pH in women. Subsequently, they studied the breakdown of whey proteins with both the female and male digestion model and found differences in proteolysis of these proteins^81^. Although the model takes into account more factors than the FGCV, it does show that the differences in gastric acid secretion between women and men are of clinical relevance to our digestion. It also highlights the importance of taking into account sex differences, both in *in vitro* digestion studies as well as *in vivo*.

In addition to the digestion of food, FGCV is also a crucial parameter in drug release and absorption of oral drugs^13,14^. The FGCV at the time of oral drug administration influences the dissolution of the drug. Especially since drugs are usually taken with a glass of water, the fasting volume will affect the pH. For example, a low initial volume will cause a less acidic environment, resulting in better solubility for acidic drugs^15^. These findings on individual variability can be used to optimize *in vitro* and *in silico* models for the development of novel drugs and dosage forms^15^.

### 6.4 Conclusion

To conclude, FGCV is highly variable and should range between 0-138 mL in healthy individuals. After correction for age, sex, and body size characteristics, intraindividual variability was 19 mL compared to 15 mL for interindividual variability. This indicates that FGCV is subject to day-to-day and within-day variation and is not a stable personal characteristic. No associations were found with age, body weight and size, within healthy, relatively young individuals who mostly had a healthy weight. Men had a ∼6 mL higher FGCV compared to women, after correction for the aforementioned factors. Differences in FGCV are expected to affect both digestion and drug dissolution. Exact implications of the observed variations, including the difference between women and men, remain to be studied further. Our results highlight the importance of considering (variations in) FGCV when studying digestion.

## Data Availability

All data produced in the present study are available upon reasonable request to the authors

## 7 Acknowledgments, authors’ contributions, funding, disclosures

The authors are grateful to all authors of the included studies.

JR, GC, PS conceptualized the study, LL collected and inputted data, JR analyzed the data and drafted the initial manuscript. GC, PS revised the manuscript. GC, PS had primary responsibility for final content. All authors critically reviewed and approved the final manuscript.

Conflict of interest: DNL has received an unrestricted educational grant from B Braun and speaker’s honoraria from Nestlé, Abbott and Corza for unrelated work. RS has received research grants from Zespri International and Sanofi and speakers fees from Ardelyx, Menarini & Ferrer. All other authors declare no conflict of interest.

Funding: No external funding was received for this study. This research was supported by the National Institute for Health Research (NIHR) Nottingham Biomedical Research Centre. The views expressed are those of the authors and not necessarily those of the NHS, the NIHR, or the Department of Health & Social Care.

## References

1. Witt T, Stokes JR. Physics of food structure breakdown and bolus formation during oral processing of hard and soft solids. Current Opinion in Food Science. 2015/06/01/ 2015;3:110-117. doi:10.1016/j.cofs.2015.06.011

2. Koç H, Vinyard CJ, Essick GK, Foegeding EA. Food Oral Processing: Conversion of Food Structure to Textural Perception. Annual Review of Food Science and Technology. 2013/02/28 2013;4(1):237-266. doi:10.1146/annurev-food-030212-182637

3. Mackie. The digestive tract: A complex system. Interdisciplinary approaches to food digestion. Springer; 2019:11–27.

4. Martinsen TC, Fossmark R, Waldum HL. The phylogeny and biological function of gastric juice— microbiological consequences of removing gastric acid. International Journal of Molecular Sciences. 2019;20(23):6031.

5. Chew CS. Gastric Acid Secretion. In: Johnson LR, ed. Encyclopedia of Gastroenterology. Elsevier; 2004:105-116.

6. Mennah-Govela YA, Swackhamer C, Bornhorst GM. Gastric secretion rate and protein concentration impact intragastric pH and protein hydrolysis during dynamic in vitro gastric digestion. Food Hydrocolloids for Health. 2021;1:100027.

7. Wilson RL, Stevenson CE. Anatomy and physiology of the stomach. Shackelford’s Surgery of the Alimentary Tract, 2 Volume Set. Elsevier; 2019:634-646.

8. Heda R, Toro F, Tombazzi CR. Physiology, Pepsin. 2019.

9. Camps G, van Eijnatten EJ, van Lieshout GA, Lambers TT, Smeets PA. Gastric Emptying and Intragastric Behavior of Breast Milk and Infant Formula in Lactating Mothers. The Journal of nutrition. 2021;151(12):3718–3724.

10. Roelofs JJM, Tjoelker RS, Lambers TT, Smeets PAM. Intragastric behavior of an experimental infant formula may better mimic intragastric behavior of human milk as compared to a control formula. medRxiv. 2023:2023.09.06.23295112. doi:10.1101/2023.09.06.23295112

11. Gardner JD, Ciociola AA, Robinson M. Measurement of meal-stimulated gastric acid secretion by in vivo gastric autotitration. Journal of Applied Physiology. 2002;92(2):427–434.

12. Pearson J, Ward R, Allen A, Roberts N, Taylor W. Mucus degradation by pepsin: comparison of mucolytic activity of human pepsin 1 and pepsin 3: implications in peptic ulceration. Gut. 1986;27(3):243.

13. van den Abeele J, Rubbens J, Brouwers J, Augustijns P. The dynamic gastric environment and its impact on drug and formulation behaviour. European Journal of Pharmaceutical Sciences. 2017;96:207–231.

14. Koziolek M, Grimm M, Schneider F, et al. Navigating the human gastrointestinal tract for oral drug delivery: Uncharted waters and new frontiers. Advanced Drug Delivery Reviews. 2016;101:75–88.

15. Grimm M, Koziolek M, Kühn J-P, Weitschies W. Interindividual and intraindividual variability of fasted state gastric fluid volume and gastric emptying of water. European Journal of Pharmaceutics and Biopharmaceutics. 2018/06/01/ 2018;127:309-317. doi:10.1016/j.ejpb.2018.03.002

16. Miraglia C, Moccia F, Russo M, et al. Non-invasive method for the assessment of gastric acid secretion. Acta Biomed. Dec 17 2018;89(8-s):53-57. doi:10.23750/abm.v89i8-S.7986

17. Goldschmiedt M, Barnett CC, Schwarz BE, Karnes WE, Redfern JS, Feldman M. Effect of age on gastric acid secretion and serum gastrin concentrations in healthy men and women. Gastroenterology. 1991;101(4):977–990.

18. Feldman M, Cryer B, McArthur KE, Huet BA, Lee E. Effects of aging and gastritis on gastric acid and pepsin secretion in humans: A prospective study. Gastroenterology. 1996/04/01/ 1996;110(4):1043-1052. doi:10.1053/gast.1996.v110.pm8612992

19. Feldman M, Barnett C. Fasting gastric pH and its relationship to true hypochlorhydria in humans. Digestive diseases and sciences. 1991;36:866–869.

20. Whitfield P, Hobsley M. Comparison of maximal gastric secretion in smokers and non-smokers with and without duodenal ulcer. Gut. 1987;28(5):557–560.

21. Baron J. Lean body mass, gastric acid, and peptic ulcer. Gut. 1969;10(8):637–642.

22. Hassan M, Hobsley M. The accurate assessment of maximal gastric secretion in control subjects and patients with duodenal ulcer. Journal of British Surgery. 1971;58(3):171–179.

23. Vakiland B, Mulekar A. Studies with the maximal histamine test. Gut. 1965;6(4):364.

24. Kekki M, Samloff I, Ihamäki T, Varis K, Siurala M. Age-and sex-related behaviour of gastric acid secretion at the population level. Scandinavian journal of gastroenterology. 1982;17(6):737–743.

25. Freire AC, Basit AW, Choudhary R, Piong CW, Merchant HA. Does sex matter? The influence of gender on gastrointestinal physiology and drug delivery. International Journal of Pharmaceutics. 2011/08/30/ 2011;415(1):15-28. doi:10.1016/j.ijpharm.2011.04.069

26. Novis B, Marks I, Bank S, Sloan A. The relation between gastric acid secretion and body habitus, blood groups, smoking, and the subsequent development of dyspepsia and duodenal ulcer. Gut. 1973;14(2):107–112.

27. Ghosh T, Lewis DI, Axon A, Everett S. Review article: methods of measuring gastric acid secretion. Alimentary pharmacology & therapeutics. 2011;33(7):768–781.

28. INFOGEST and UNGAP Imaging Working Group. https://www.gastrointestinalmri.org.uk/infogest-ungap-imaging-working-group/

29. Sheather S. A modern approach to regression with R. Springer Science & Business Media; 2009.

30. Schielzeth H, Dingemanse NJ, Nakagawa S, et al. Robustness of linear mixed-effects models to violations of distributional assumptions. Methods in ecology and evolution. 2020;11(9):1141–1152.

31. Pinheiro J, Bates D, DebRoy S, et al. Package ‘nlme’. Linear and nonlinear mixed effects models, version. 2017;3(1):274.

32. Fox J, Weisberg S, Adler D, et al. Package ‘car’. Vienna: R Foundation for Statistical Computing. 2012;16

33. ClinicalTrials.gov NCT05575687. Unpublished.

34. ClinicalTrials.gov NCT05854407. Unpublished.

35. trialsearch.who.int NL8137. Unpublished.

36. Murray K, Placidi E, Schuring EAH, et al. Aerated drinks increase gastric volume and reduce appetite as assessed by MRI: a randomized, balanced, crossover trial234. The American Journal of Clinical Nutrition. 2015/02/01/ 2015;101(2):270-278. doi:10.3945/ajcn.114.096974

37. Alyami J, Whitehouse E, Yakubov GE, et al. Glycaemic, gastrointestinal, hormonal and appetitive responses to pearl millet or oats porridge breakfasts: a randomised, crossover trial in healthy humans. British Journal of Nutrition. 2019;122(10):1142–1154. doi:10.1017/S0007114519001880

38. Freitas D, Boué F, Benallaoua M, et al. Glycemic response, satiety, gastric secretions and emptying after bread consumption with water, tea or lemon juice: a randomized crossover intervention using MRI. European Journal of Nutrition. 2022/04/01 2022;61(3):1621-1636. doi:10.1007/s00394-021-02762-2

39. Boulby P, Gowland P, Adams V, Spiller R. Use of echo planar imaging to demonstrate the effect of posture on the intragastric distribution and emptying of an oil/water meal. Neurogastroenterology & Motility. 1997;9(1):41–47.

40. Grimm M, Koziolek M, Saleh M, et al. Gastric Emptying and Small Bowel Water Content after Administration of Grapefruit Juice Compared to Water and Isocaloric Solutions of Glucose and Fructose: A Four-Way Crossover MRI Pilot Study in Healthy Subjects. Molecular Pharmaceutics. 2018/02/05 2018;15(2):548-559. doi:10.1021/acs.molpharmaceut.7b00919

41. Mudie DM, Murray K, Hoad CL, et al. Quantification of Gastrointestinal Liquid Volumes and Distribution Following a 240 mL Dose of Water in the Fasted State. Molecular Pharmaceutics. 2014/09/02 2014;11(9):3039-3047. doi:10.1021/mp500210c

42. Krishnasamy S, Lomer MCE, Marciani L, et al. Processing Apples to Puree or Juice Speeds Gastric Emptying and Reduces Postprandial Intestinal Volumes and Satiety in Healthy Adults. The Journal of Nutrition. 2020/11/01/ 2020;150(11):2890-2899. doi:10.1093/jn/nxaa191

43. Camps G, Veit R, Mars M, de Graaf C, Smeets PA. Just add water: Effects of added gastric distention by water on gastric emptying and satiety related brain activity. Appetite. 2018;127:195–202.

44. Coletta M, Gates FK, Marciani L, et al. Effect of bread gluten content on gastrointestinal function: a crossover MRI study on healthy humans. British Journal of Nutrition. 2016;115(1):55–61. doi:10.1017/S0007114515004183

45. Deng R, Mars M, Janssen AEM, Smeets PAM. Gastric digestion of whey protein gels: A randomized cross-over trial with the use of MRI. Food Hydrocolloids. 2023/08/01/ 2023;141:108689. doi:10.1016/j.foodhyd.2023.108689

46. Hussein MO, Hoad CL, Wright J, et al. Fat Emulsion Intragastric Stability and Droplet Size Modulate Gastrointestinal Responses and Subsequent Food Intake in Young AdultsNitrogen1, 2, 3, 4. The Journal of Nutrition. 2015/06/01/ 2015;145(6):1170-1177. doi:10.3945/jn.114.204339

47. Juvonen KR, Macierzanka A, Lille ME, et al. Cross-linking of sodium caseinate-structured emulsion with transglutaminase alters postprandial metabolic and appetite responses in healthy young individuals. British Journal of Nutrition. 2015;114(3):418–429. doi:10.1017/S0007114515001737

48. Lobo DN, Hendry PO, Rodrigues G, et al. Gastric emptying of three liquid oral preoperative metabolic preconditioning regimens measured by magnetic resonance imaging in healthy adult volunteers: A randomised double-blind, crossover study. Clinical Nutrition. 2009/12/01/ 2009;28(6):636-641. doi:10.1016/j.clnu.2009.05.002

49. Marciani L, Cox EF, Hoad CL, et al. Postprandial Changes in Small Bowel Water Content in Healthy Subjects and Patients With Irritable Bowel Syndrome. Gastroenterology. 2010/02/01/ 2010;138(2):469-477.e1. doi:10.1053/j.gastro.2009.10.055

50. Marciani L, Hall N, Pritchard SE, et al. Preventing Gastric Sieving by Blending a Solid/Water Meal Enhances Satiation in Healthy Humans. The Journal of Nutrition. 2012/07/01/ 2012;142(7):1253-1258. doi:10.3945/jn.112.159830

51. Marciani L, Pritchard SE, Hellier-Woods C, et al. Delayed gastric emptying and reduced postprandial small bowel water content of equicaloric whole meal bread versus rice meals in healthy subjects: novel MRI insights. European Journal of Clinical Nutrition. 2013/07/01 2013;67(7):754-758. doi:10.1038/ejcn.2013.78

52. Marciani L, Cox EF, Pritchard SE, et al. Additive effects of gastric volumes and macronutrient composition on the sensation of postprandial fullness in humans. European Journal of Clinical Nutrition. 2015/03/01 2015;69(3):380-384. doi:10.1038/ejcn.2014.194

53. Roelofs JJM, van Eijnatten EJM, Prathumars P, et al. Gastric emptying and nutrient absorption of pea protein products differing in heat treatment and texture: A randomized in vivo crossover trial and in vitro digestion study. Food Hydrocolloids. 2024/04/01/ 2024;149:109596. doi:10.1016/j.foodhyd.2023.109596

54. van Eijnatten EJM, Roelofs JJM, Camps G, Huppertz T, Lambers TT, Smeets PAM. Gastric coagulation and postprandial amino acid absorption of milk is affected by mineral composition: a randomized crossover trial. medRxiv. 2023:2023.09.13.23295475. doi:10.1101/2023.09.13.23295475

55. van Eijnatten EJ, Camps G, Guerville M, Fogliano V, Hettinga K, Smeets PAM. Milk coagulation and gastric emptying in women experiencing gastrointestinal symptoms after ingestion of cow’s milk. Neurogastroenterology & Motility. 2023;36(1):e14696. doi:10.1111/nmo.14696

56. Vantrappen G, Peeters T, Janssens J. The secretory component of the interdigestive migrating motor complex in man. Scandinavian journal of gastroenterology. 1979;14(6):663–667.

57. Parkman HP, Urbain JL, Knight LC, et al. Effect of gastric acid suppressants on human gastric motility. Gut. Feb 1998;42(2):243–50. doi:10.1136/gut.42.2.243

58. Kellow J, Borody T, Phillips S, Tucker R, Haddad A. Human interdigestive motility: variations in patterns from esophagus to colon. Gastroenterology. 1986;91(2):386–395.

59. Goetze O, Treier R, Fox M, et al. The effect of gastric secretion on gastric physiology and emptying in the fasted and fed state assessed by magnetic resonance imaging. Neurogastroenterology & Motility. 2009;21(7):725–e42.

60. Goyal R, Gupta P, Chuttani K. Gastric acid secretion in Indians with particular reference to the ratio of basal to maximal acid output. Gut. 1966;7(6):619.

61. Rahim Z, Yaacob H. Effects of fasting on saliva composition. The Journal of Nihon University School of Dentistry. 1991;33(4):205–210.

62. Acosta A, Camilleri M, Shin A, et al. Quantitative Gastrointestinal and Psychological Traits Associated With Obesity and Response to Weight-Loss Therapy. Gastroenterology. 2015/03/01/ 2015;148(3):537-546.e4. 10.1053/j.gastro.2014.11.020

63. Cox A. Variations in size of the human stomach. California and western medicine. 1945;63(6):267.

64. Lee E-G, Kim T-H, Huh Y-J, et al. Anthropometric study of the stomach. Journal of Gastric Cancer. 2016;16(4):247–253.

65. Sakaguchi T, Yamazaki M, Itoh S, Okamura N, Bando T. Gastric acid secretion controlled by oestrogen in women. Journal of international medical research. 1991;19(5):384–388.

66. Campbell-Thompson M, Reyher KK, Wilkinson L. Immunolocalization of estrogen receptor alpha and beta in gastric epithelium and enteric neurons. Journal of endocrinology. 2001;171(1):65–73.

67. Moore J, Englert E. Circadian rhythm of gastric acid secretion in man. Nature. 1970;226(5252)

68. Eurostat. Daily smokers of cigarettes by sex, age and educational attainment level. Updated 04/04/2022. Accessed 31/01, 2024. https://ec.europa.eu/eurostat/databrowser/view/hlth_ehis_sk3e__custom_9586928/default/table?lang=en&page=time:2019

69. Zach E, Markiewicz K, Lukin M, Cholewa M. [The behaviour of basal gastric secretion during exercise and restitution in chronic gastric and duodenal ulcer patients (author’s transl)]. Dtsch Z Verdau Stoffwechselkr. 1982 1982;42(2-3):53–63.

70. Markiewicz K, Cholewa M, Górski L, Chmura J. Effect of physical exercise on gastric basal secretion in healthy men. Acta Hepatogastroenterol (Stuttg). 1977/10// 1977;24(5):377-380.

71. Canelles P, Diago M, Tomé A, Medina E, Orti E, Martínez A. [Physical exercise and gastric acid secretion]. Rev Esp Enferm Dig. 1990/03// 1990;77(3):179-184.

72. Oektedalen O, Guldvog I, Opstad P, Berstad A, Gedde-Dahl D, Jorde R. The effect of physical stress on gastric secretion and pancreatic polypeptide levels in man. Scandinavian journal of gastroenterology. 1984;19(6):770–778.

73. Sanaka M, Yamamoto T, Kuyama Y. Effects of proton pump inhibitors on gastric emptying: a systematic review. Dig Dis Sci. Sep 2010;55(9):2431–40. doi:10.1007/s10620-009-1076-x

74. Maideen NMP. Adverse Effects Associated with Long-Term Use of Proton Pump Inhibitors. Chonnam Med J. May 2023;59(2):115–127. doi:10.4068/cmj.2023.59.2.115

75. Calam J. Helicobacter pylori modulation of gastric acid. Yale J Biol Med. Mar-Jun 1999;72(2-3):195–202.

76. Hooi JKY, Lai WY, Ng WK, et al. Global Prevalence of Helicobacter pylori Infection: Systematic Review and Meta-Analysis. Gastroenterology. 2017/08/01/ 2017;153(2):420-429. 10.1053/j.gastro.2017.04.022

77. Feldman M, Richardson CT. Role of thought, sight, smell, and taste of food in the cephalic phase of gastric acid secretion in humans. Gastroenterology. 1986;90(2):428–433.

78. Lennernäs H. Ethanol− drug absorption interaction: Potential for a significant effect on the plasma pharmacokinetics of ethanol vulnerable formulations. Molecular pharmaceutics. 2009;6(5):1429–1440.

79. Varum FJO, Hatton GB, Basit AW. Food, physiology and drug delivery. International Journal of Pharmaceutics. 2013/12/05/ 2013;457(2):446-460. doi:10.1016/j.ijpharm.2013.04.034

80. Marciani L, Gowland PA, Spiller RC, et al. Effect of meal viscosity and nutrients on satiety, intragastric dilution, and emptying assessed by MRI. American Journal of Physiology-Gastrointestinal and Liver Physiology. 2001;280(6):G1227–G1233.

81. Lajterer C, Levi CS, Lesmes U. An in vitro digestion model accounting for sex differences in gastro-intestinal functions and its application to study differential protein digestibility. Food Hydrocolloids. 2022;132:107850.

