## Supplementary materials for "Intra- and interindividual variability in fasted gastric content volume"

### Supplement

#### Supplementary Tables

Supplementary Table 1. Fasting instructions for the participants of each study.

| Study | Fasting | Drinking | Exercise | Medication | Meal |
|---|---|---|---|---|---|
| **Alyami et al.[37]** | >10h | No drinking, only one glass upon waking | | Excluded | |
| **Camps et al.[43]** | Overnight | | | Excluded | |
| **Camps et al.[9]** | >10h | Water allowed up to 1h before test session | | Excluded | |
| **ClinicalTrials.gov NCT05575687[33]** | After 20PM | Water allowed up to 1h before test session | | Excluded | |
| **ClinicalTrials.gov NCT05854407[34]** | After 22PM | Water allowed up to 1,5h before test session | | Excluded | Same meal |
| **Coletta et al.[44]** | >12h | No drinking, only small glass upon waking | 18h strenuous exercise | 18h | |
| **Deng et al.[45]** | After 22PM | Water allowed up to 1,5h before test session | | Excluded | |
| **Freitas et al.[38]** | >10h | | No excessive PA evening before | Excluded | |
| **Grimm et al.[40]** | >10h | | | Stopped during study | |
| **Hussein et al.[46]** | After 20PM | | 18h | 24h | |
| **Juvonen et al.[47]** | 10-12h | | 24h no heavy exercise | Excluded | |
| **Krishnasamy et al.[42]** | After 22PM | No drinking, only small glass (50mL) upon waking | 18h strenuous exercise | Stopped during study | |
| **Lobo et al.[48]** | Overnight | | 18h strenuous exercise | 24h | |
| **Marciani et al.[49]** | Overnight | Not allowed | overnight | Overnight | |
| **Marciani et al.[50]** | >12h | | 18h | 18h | |
| **Marciani et al.[51]** | >13h | No drinking, only one glass upon waking | 18h | 18h | |
| **Marciani et al.[52]** | After evening meal | | 18h | 24h | light, non-fatty meal |
| **Mudie et al.[41]** | >10h | Water allowed up to 1h before test session | | Excluded | Same size as usual |

| | | | | | |
|---|---|---|---|---|---|
| **Murray et al.[36]** | After 22PM | Water allowed before arrival | 24h | Excluded | |
| **Roelofs et al.[10]** | >12h | Water allowed up to 2h before test session | Keep constant for both days | Excluded | Same meal |
| **Roelofs et al.[53]** | >12h | Water allowed up to 1,5h before test session | Keep constant for both days | Excluded | Standardized meal |
| **Trialsearch.who.int NL8137[35]** | >12h | Water allowed up to 1,5h before test session | Keep constant for both days | Excluded | |
| **van Eijnatten et al.[54]** | >12h | Water and herbal tea allowed up to 1,5h before test session | | Excluded | Standardized meal |
| **van Eijnatten et al.[55]** | After 20PM | Non-caloric, non-caffeinated liquids like water or herbal tea up to 1h before test session | | Excluded | |

Supplemental Table 2. In- and exclusion criteria.

| Study | Inclusion criteria: Sex | Age | BMI | Controlled for: GI related medication | GI surgery | GI disorders | Alcohol | Smoking | Recreational drugs | Weight gain/loss |
|---|---|---|---|---|---|---|---|---|---|---|
| **Alyami et al.[37]** | F/M | 18-65 | 18-25 & <120kg | Yes | Yes | Yes | >21 units/w | | | >10% in last 6 months |
| **Camps et al.[43]** | M | 18-35 | 18-25 | | Yes | Yes | | | | >5 kg in last 2 months |
| **Camps et al.[9]** | F | None | None | Yes | | Yes | | Yes | | |
| **ClinicalTrials.gov NCT05575687[33]** | F/M | 18-45 | 18,5-25 | Yes | Yes | Yes | >7 units/w | >2/w | <1 w | >5kg in last month |
| **ClinicalTrials.gov NCT05854407[34]** | F/M | 18-30 | 18,5-25 | Yes | Yes | Yes | >14 units/w | >1/d | <1 w | |
| **Coletta et al.[44]** | F/M | 18-55 | <120 kg | Discontinued 2 weeks prior to study | Yes | Yes | Dependence | Discontinued during study | | |
| **Deng et al.[45]** | M | 18-45 | 18.5-25 | Yes | Yes | Yes | | | | |
| **Freitas et al.[38]** | M | 18-60 | 18-25 | | Yes | Yes | Abusive consumption | If started/stopped in <3 months | | >3 kg in last 3 months |
| **Grimm et al.[40]** | F/M | | | Discontinued during study | | | Discontinued during study | | | |
| **Hussein et al.[46]** | F/M | | | | | | | | | |
| **Juvonen et al.[47]** | F/M | | Normal | Yes | | | >21 units/w | Yes | | |
| **Krishnasamy et al.[42]** | F/M | 18+ | <120 kg | Discontinued during study | Yes | Yes | >35 units/w or 8 units/d | No | | |
| **Lobo et al.[48]** | F/M | 18-45 | 20-26 | | | | | | | |
| **Marciani et al.[49]** | F/M | 21-28 | | | | | | | | |
| **Marciani et al.[50]** | F/M | | Normal | | | | | | | |

| | | | | | | | | | | | | |
|---|---|---|---|---|---|---|---|---|---|---|---|---|
| **Marciani et al.[51]** | F/M | | | | | | | | | | | |
| **Marciani et al.[52]** | F/M | | | | | | | | | | | |
| **Mudie et al.[41]** | F/M | 18-55 | 18,5-25 | Yes | | Yes | Yes | >21 units/w | Yes | | If history of abuse | |
| **Murray et al.[36]** | M | 18-60 | 20-35 | Yes | | Yes | Yes | >21 units/w | Yes | | | >10% in last 6 months |
| **Roelofs et al.[10]** | M | 18-45 | 18,5-25 | Yes | | Yes | Yes | >14 units/w | >2/w | | | >5 kg in last 2 months |
| **Roelofs et al.[53]** | M | 18-55 | 18,5-25 | Yes | | Yes | Yes | >14 units/w | Yes | | <4 w | >5 kg in last month |
| **Trialsearch.who.int NL8137[35]** | M | 18-55 | 18-25 | Yes | | | Yes | | Yes | | | |
| **van Eijnatten et al.[54]** | M | 18-55 | 18,5-25 | Yes | | Yes | Yes | >14 units/w | Yes | | <4 w | >5 kg in last 2 months |
| **van Eijnatten et al.[55]** | F | 18-60 | 18,5-30 | Yes | | Yes | Yes | >14 units/w | >4/d | | | >5kg in last month |

Supplementary Table 3. MRI details.

| Study | Magnetic Field (T) | Type of scan sequence | # Slices | Resolution* (mm) | Interslice gap (mm) |
|---|---|---|---|---|---|
| **Alyami et al.[37]** | 1.5 | Balanced turbo field echo | 25 | | |
| **Camps et al.[43]** | 3.0 | Turbo spin-echo | 24 | 1.19*1.19*6 | 2.4 |
| **Camps et al.[9]** | 3.0 | Turbo spin-echo | 24 | 1.19*1.19*6 | 2.4 |
| **ClinicalTrials.gov NCT05575687[33]** | 3.0 | 2-D Turbo Spin Echo | 33 | 1.00*1.00*5 | 1.4 |
| **ClinicalTrials.gov NCT05854407[34]** | 3.0 | Turbo Spin Echo | 28 | 0.625*0.625*4 | 1.4 |
| **Coletta et al.[44]** | 1.5 | Balanced turbo field echo | 25 | 2.01*1.76*10 | 0 |
| **Deng et al.[45]** | 1.5 | Spin-echo | 33 | 0.78*0.78*6 | |
| **Freitas et al.[38]** | 1.5 | Turbo spin-echo | 72 | 1.67*1.71*3 | 0 |
| **Grimm et al.[40]** | 1.5 | Turbo spin-echo | 40** | 1.76*1.7*5.1 | 0.77 |
| **Hussein et al.[46]** | 1.5 | Balanced turbo field echo | 40 | 1.56*1.56*7 | 0 |
| **Juvonen et al.[47]** | 1.5 | Balanced fast field echo | | 1.56*1.56*5 | 0 |
| **Krishnasamy et al.[42]** | 1.5 | Balanced turbo field echo | 50 | 1.56*1.56*5 | 0 |
| **Lobo et al.[48]** | 1.5 | Balanced turbo field echo | 40 | 2.5*1.56*10 | |
| **Marciani et al.[49]** | 1.5 | Balanced turbo field echo | 20 | 1.56*1.56*10 | 0 |
| **Marciani et al.[50]** | 1.5 | Balanced turbo field echo | 24 | 1.56*1.56*10 | 0 |
| **Marciani et al.[51]** | 1.5 | Balanced turbo field echo | 20 | 1.56*1.56*10 | 0 |
| **Marciani et al.[52]** | 1.5 | Balanced turbo field echo | 24 | 1.56*1.56*10 | 0 |
| **Mudie et al.[41]** | 1.5 | Balanced turbo field echo | 50 | 2.00*1.77*5 | 0 |
| **Murray et al.[36]** | 1.5 | Single-shot balanced-gradient echo | 30 | 1.56*1.57*10 | 0 |
| **Roelofs et al.[10]** | 3.0 | 2-D Turbo Spin Echo | 37 | 1.00*1.00*4 | 1.4 |
| **Roelofs et al.[53]** | 3.0 | 2-D Turbo Spin Echo | 37 | 1.00*1.00*4 | 1.4 |
| **Trialsearch.who.int NL8137[35]** | 3.0 | Turbo spin-echo | 24 | 1.19*1.19*6 | 2.4 |
| **van Eijnatten et al.[54]** | 3.0 | 2-D Turbo Spin Echo | 37 | 1.00*1.00*4 | 2 |
| **van Eijnatten et al.[55]** | 3.0 | Turbo spin-echo | 24 | 1.19*1.19*6 | 2.4 |

*Without gap; **coronal slices

Supplementary Table 4. Mean fasted gastric content volume and variability with outlier.

| | |
|---|---|
| **Mean** | 33 mL |
| **Median** | 27 mL |
| **Range** | 0.0 – 373 mL |
| **Coefficient of variation** | 82.5% |
| **Interindividual variability** | 14 mL |
| **Intraindividual variability** | 23 mL |

Supplementary Table 5. Results of the linear mixed model analysis of the effect of age, sex, and body size characteristics on fasted gastric content volume (mL unit$^{-1}$) with outlier.

| | **Age, Sex, Weight & Height** | | **Age, Sex, Weight*Height & BMI** | |
|---|---|---|---|---|
| | *Estimate* | *p-value* | *Estimate* | *p-value* |
| **Age** | 0 mL y$^{-1}$ | 0.205 | 0 y$^{-1}$ | 0.189 |
| **Sex** | -7 mL | 0.027 | -7 mL | 0.024 |
| **Weight** | 0 mL kg$^{-1}$ | 0.796 | | |
| **Height** | 0 mL m$^{-1}$ | 0.261 | | |
| **Weight*Height** | | | 0 mL (kg*m)$^{-1}$ | 0.186 |
| **BMI** | | | 1 mL (kg m$^{-2}$)$^{-1}$ | 0.259 |

Supplementary Figure

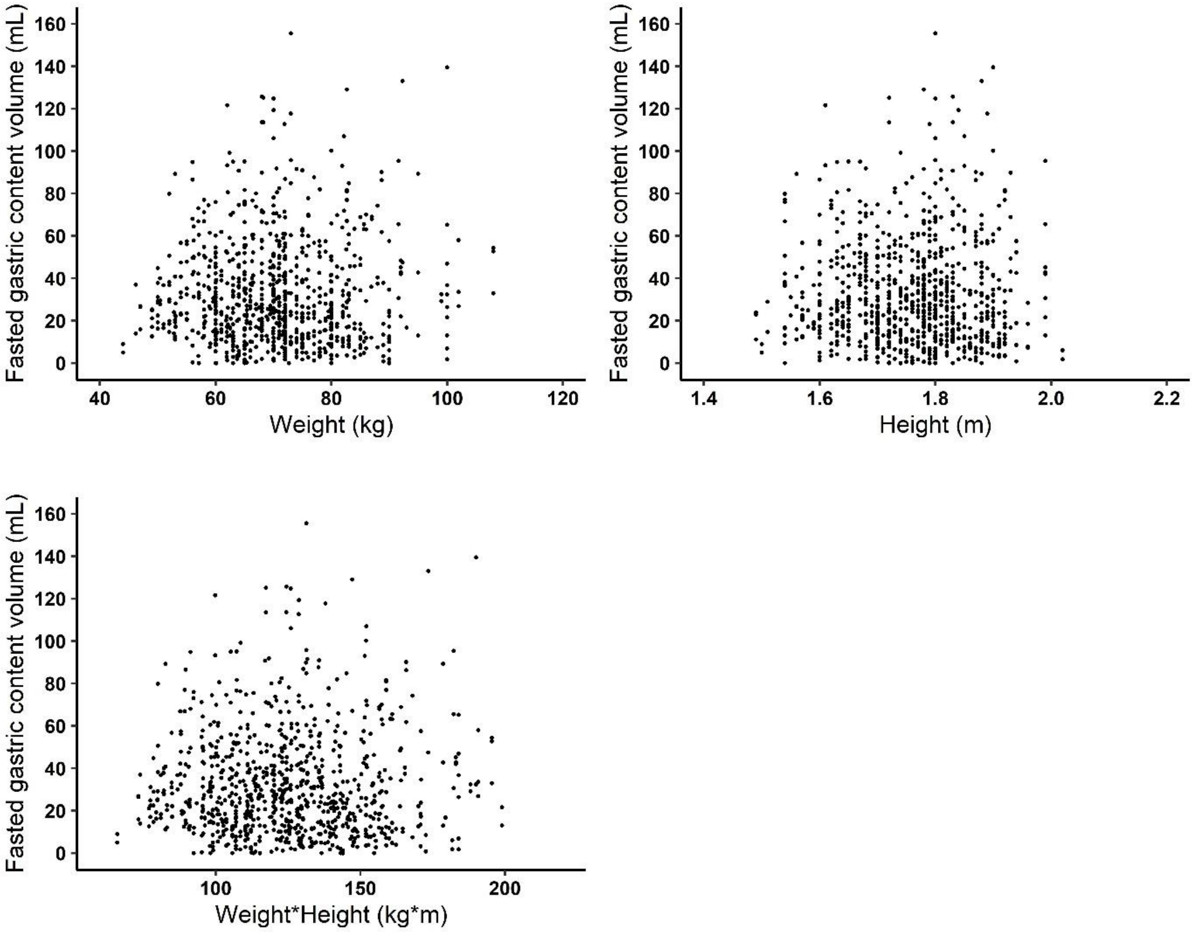

Supplementary Figure 1. Scatterplots of fasted gastric content volume by weight, height, and weight*height (all $p > 0.05$).
